# Superspreading of SARS-CoV-2 infections: A Systematic Review and Meta-analysis

**DOI:** 10.1101/2021.12.09.21267507

**Authors:** Zhanwei Du, Chunyu Wang, Caifen Liu, Yuan Bai, Sen Pei, Dillon C. Adam, Lin Wang, Peng Wu, Eric H. Y. Lau, Benjamin J. Cowling

## Abstract

Superspreading in transmission is a feature of SARS-CoV-2 transmission. We conducted a systematic review and meta-analysis on globally reported dispersion parameters of SARS-CoV-2. The pooled estimate was 0.55 (95% CI: 0.30, 0.79). The study location and method were found to be important drivers for its diversity.

## Main Text

A novel coronavirus (SARS-CoV-2) was first identified in Wuhan, China, in early 2020 and rapidly spread throughout the world. The World Health Organization (WHO) declared a pandemic on March 11, 2020 [1]. As of October 12, 2021, over 219 million confirmed COVID-19 cases and 4.55 million deaths have been reported [2]. Worldwide, four variants of concern (VOC) and two variants of interest (VOI) have already been identified by WHO to-date [3]. Some of these variants have exhibited increased transmissibility and severity compared to wild-type SARS-CoV-2 virus; with some also able to partially evade immunity conferred by prior infection or vaccination [4].

The dispersion parameter (*k*) is a statistical parameter used to characterize and quantify heterogeneity in certain distributions. In the context of measuring transmissibility, overdispersion in transmission has often been estimated by assuming that the collective offspring distribution follows a negative-binomial distribution [5]. Specifically, the variance of the number of secondary infections from each case is *R*+*R*^2^/*k*, where *R* is the mean and *k* is the dispersion parameter. A small value of *k* indicates increased heterogeneity in transmission and therefore a high potential of superspreading, and describes the phenomenon that a few infectious cases account for most secondary transmissions. Accurate estimates of *k* are essential for determining the potential need for, and intensity of, public health and social measures (PHSMs) needed for disease control. When superspreading potential is low, i.e., *k* is high, relaxing PHSMs to reopen societies becomes feasible. While under high potential of superspreading, larger outbreaks could still occur even when an epidemic seems to be under control.

For SARS and MERS, most infections are caused by a small proportion of cases, with the dispersion parameter ranging from 0.06 to 2.94 [6]. However, a comprehensive review and comparison of the superspreading potential of COVID-19 and its uncertainty over countries is still lacking. We carried out a systematic review and meta-analysis of published estimates of the dispersion parameter, aiming to estimate the pooled *k* of SARS-CoV-2 infections.

## Methods

### Search Strategy and Selection Criteria

All searches were carried out on 10 September 2021 in PubMed for articles published from 1 January 2020 to 10 September 2021. We included all relevant articles that were published in peer reviewed journals, coupled with 8 articles recommended by experts. Search terms for superspreading for COVID-19 variants included (#1) “COVID-19” OR “SARS-COV-2” OR “2019-nCov” OR “Coronavirus 2019” OR “2019 coronavirus” OR “Wuhan coronavirus” OR “Wuhan pneumonia”; and (#2) “Superspreader” OR “Spreader” OR “Superspreader event” OR “Super-spreader” OR “Super-spreader hosts” OR “Super-spreading” OR “Superspreading” OR “Overdispersion” OR “Dispersion parameter” OR “20/80 rule” AND “dispersion parameter” and the final search term was #1 AND #2. After reading the abstract and full text, we included studies in which estimates of the dispersion parameter were reported along with their uncertainty intervals and estimation periods. We excluded other systematic reviews and meta-analysis from our analyses but included relevant studies mentioned in these reviews. Finally, 144 studies are included with the publish date between 20 March 2020 and 3 September 2021.

### Data Extraction

All data were extracted independently and entered in a standardized form by 2 co-authors (C. W. and C. L.). Conflicts over inclusion of the studies and retrieving the estimates of these variables were resolved by another co-author (Z. D.). Information was extracted on the estimates of dispersion parameters of COVID-19 superspreading coupled with the corresponding 95% or 90% confidence interval (CI) or the 95% credible interval (CrI) or 95% range across 500 instances of reconstructed transmission tree (95% Range). This paper converts 90% CI to 95% CI for meta-analysis. Other information such as study’s information (i.e., estimation period and location), model used in estimation measurements of transmissibility and heterogeneity (i.e., dispersion parameter, ‘20/80’ rule and dispersion parameter), and study population and settings (i.e., type of cases) was also extracted for each selected study (see Supplementary Materials for details).

### Estimation of dispersion parameter in studies reporting the ‘20/80′ rule

A framework is proposed to compute the dispersion parameter (*k*) by reported reproduction number (*R*) and the transmission distribution profiles in the form of the ‘20/80’ rule [5,7]. For those articles without *k* reported, we adopted the framework below to estimate *k* in Eq. (1).

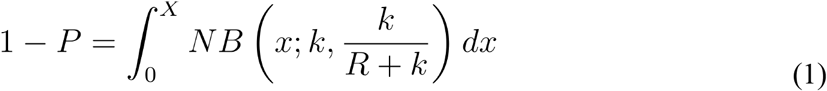

where *X* satisfies,

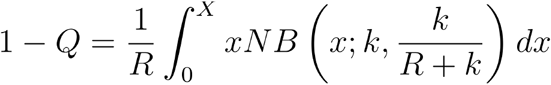

where *P* is the expected proportion of the most infectious individuals responsible for *Q* of all transmissions. *NB*(•) means the negative binomial distribution for secondary cases with mean *R* and *k*. Thus, 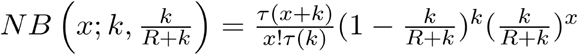, where *τ*(*k*)=(*k*−1)!.

### Statistical Analysis

We use the *I*^2^ index to assess heterogeneity between studies into the following 3 categories: *I*^*2*^<25% (low heterogeneity), *I*^*2*^=25-75% (average heterogeneity), and *I*^*2*^>75% (high heterogeneity). Because of the high *I*^*2*^ value that was calculated in our results, as well as the significance of the Cochran *Q* test, a random-effects model was further used to perform a meta-analysis in this study. Finally, meta-regression analysis using a mixed-effects model was conducted to quantify the association between study’s location and the estimate of dispersion parameter. Analyses were conducted in R version 4.1.1.

## Result

We identified 114 studies by searching PubMed and additionally included 8 studies from our own reference list. Of these, 59 studies were excluded through title and abstract screening, leaving 55 studies for full-text assessment. A total of 19 of them were finally included in this study, providing 45 estimates. The detailed selection process is illustrated in **Figure S1**. The reports are conducted based on data in 8 countries (e.g., China, USA, India, Indonesia, Israel, Japan, New Zealand, and Singapore) using 3 methods (e.g., negative binomial distribution, Zero-truncated negative binomial distribution, and phylodynamic analysis) (**Table S1**). There was no published estimate of the dispersion parameter based on data in 2021.

High heterogeneity was reported among the included studies (*I*^*2*^=100% and p<0.0001). The mean estimates of dispersion parameter (*k*) range from 0.06 to 2.97 over 8 countries. The pooled estimate of *k* was 0.55 (95% CI: 0.30, 0.79), with changing means over countries (**Figure 1**) and decreasing slightly with the increasing reproduction number (**Figure S2**). The global estimates are 0.54 (95% CI: 0.54, 8.18) in January 2020 [8] and 0.10 (95% CI: 0.05, 0.20) in February 2020 [7]. The expected proportion of cases accounting for 80% infections is 19% (95% CrI: 7, 34) over countries (**Table S2**).

**Figure 1.**
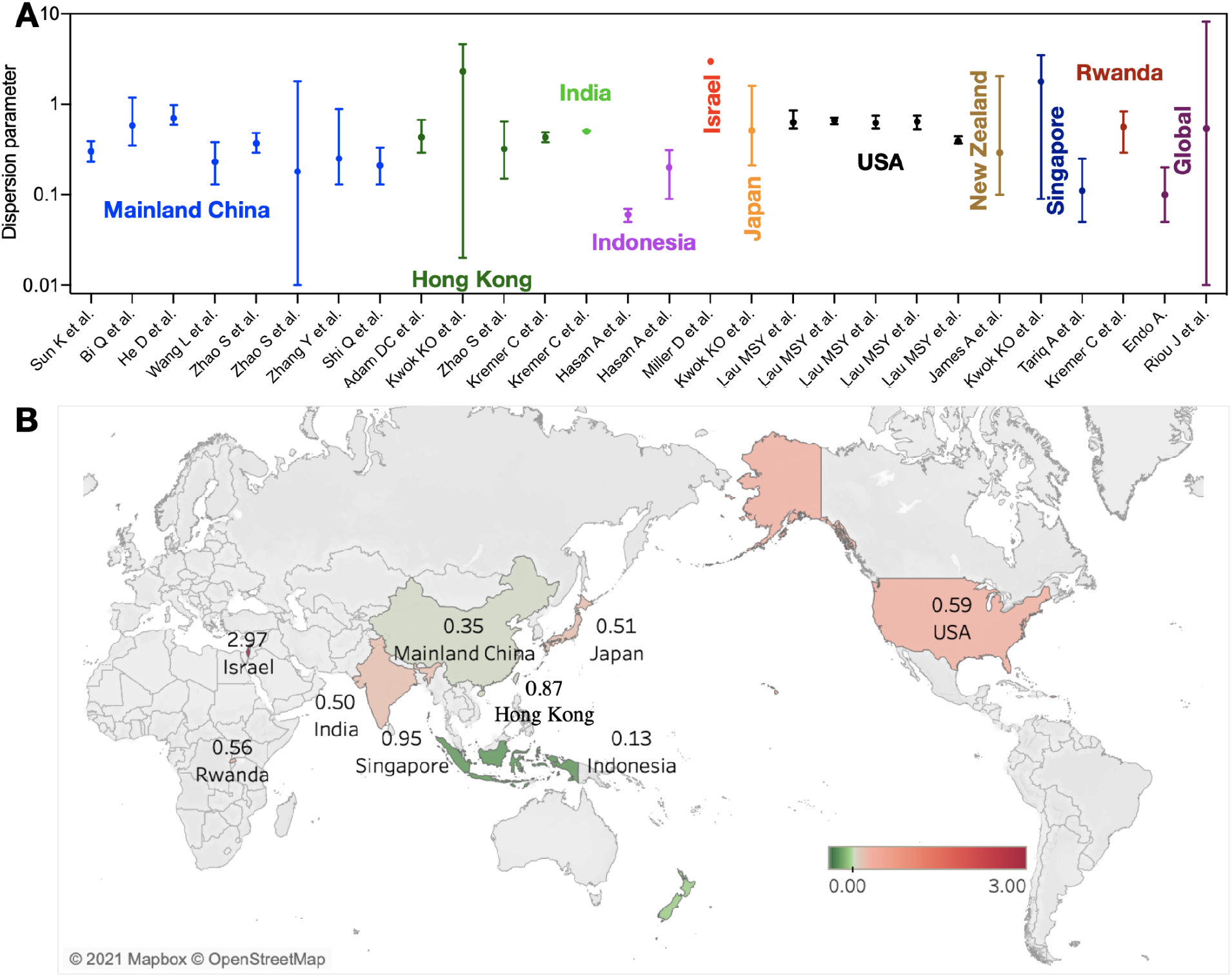
Dispersion parameter estimates for coronavirus disease 2019 (COVID-19) reported in 19 unique studies presented by country. **(A)** Estimates of dispersion parameters over countries. The error bars show the mean values and 95% confidence interval. **(B)** Mean estimate of dispersion parameters by countries over studies.

The meta-regression analysis was conducted based on the reported *k* estimates, which allowed us to explore the potential association between the study attribute (e.g., location, methods, or age groups) and the estimated dispersion parameter (**Figure S3**). We found that the study location was closely associated with the reported dispersion parameter in the meta-analysis by including country, age group, or method as a categorical variable (p<0.0001).

## Discussion

For SARS-CoV-1, SARS-CoV-2 and MERS-CoV, most infections are caused by a small proportion of people. During the 2003 SARS epidemic, 76 infections arose from 1 hospitalized patient in Beijing, China [9]. And during the 2015 MERS outbreak, 5 patients led to 154 secondary infections in South Korea [10]. In this early COVID-19 outbreak, around 10% of cases in countries outside China accounted for 80% of secondary cases [7]. But epidemiological population-level measures (e.g., the basic reproduction number) usually hides immense variation at the individual level. We thus carried out a systematic review and meta-analysis of 17 studies on the dispersion parameter to characterize COVID-19 superspreading.

Estimation of the dispersion parameter from individual case data requires accurate observation of transmission chains, usually collected through contact-tracing or phylodynamic analysis, and can be biased, perhaps by reporting bias, estimation methods and transmission scenarios. The negative binomial model with the zero-truncated framework would reduce the estimation bias of dispersion parameter when the under-ascertainment of index cases with zero secondary case occurs, for example in China [11]. Estimating and monitoring changes in the dispersion parameter is thus critical for determining the type and stringency of public health and social measures (PHSMs) needed to reduce the occurrence of superspreading events, although we found that the estimate for the variant Delta or even any other variant is not yet available. Japan recognized the importance of superspreading in February 2020, implemented the cluster-focused backwards contact tracing, and promoted awareness of people at risk of infection by closing higher risk locations, followed by the World Health Organization’s Western Pacific Region in July 2020 to limit the number of people to gather indoors thus to curb the spread of the virus. And restaurants were estimated to account for 20% of transmissions if all businesses were to reopen in 2020 in the USA [12]. Such measures can mitigate the impact of superspreading events, which are expected to be major drivers in early epidemics.

In the recent systematic review of COVID-19 superspreading by 10 February, 2021, the estimates of dispersion parameters for COVID-19 range from 0.01 in the United States to 5 in Israel [6]. We include most of their studies together with those published by 10 September 2021, and re-estimate those based on some simple assumptions to conduct the pooled estimates and the meta-analysis. The major difference is the lower dispersion parameter, which is estimated to be 0.01 in the United States in the published review [6]. In contrast, we directly extract the estimates from figures in the raw study, which range from 0.39 to 0.66 before the shelter-in-place order, resulting in the lower limit changing to 0.06 as that in Indonesia (**Table S1**). Finally, the pooled estimates from our analysis indicated that the dispersion parameter of COVID-19 was likely to be 0.55 (95% CI: 0.30, 0.79), approximate to that of India, China, and USA (**Figure 1**).

Our study has several limitations. Most articles included in our study used publicly available data. Some studies in our review might have used overlapping data, leading to double counting in the pooled estimates. And with the recent emergence of variants that may be more transmissible and evade immunity acquired through prior infection or vaccination, the future of the pandemic is highly uncertain. Meanwhile, SARS-CoV-2 viruses are constantly evolving through mutation; genetic variations have emerged and circulated over the world which may modify individual infectiousness profiles. We are still not clear about the impact of variants on overdispersion, perhaps by increasing transmissibility. Our pooled estimate is based on the previous transmission of wide-type in early 2020, which may not be generalisable to the dominant variant Delta and future studies will be needed to conduct the comparison.

In conclusion, multiple estimates of the dispersion parameter have been published for 17 studies, which could be related to where and when the data was obtained. The study location and method were found to be important drivers for diversity in estimates of dispersion parameters.

## Data Availability

Not applicable.

## Acknowledgments

We acknowledge the financial support from the Collaborative Research Fund (Project No. C7123-20G) of the Research Grants Council of the Hong Kong SAR Government.

## Author Contributions

ZW, CW, CL and BJC: conceived the study, designed statistical and modelling methods, conducted analyses, interpreted results, wrote and revised the manuscript; YB, SP, DA, LW, PW, and EL: interpreted results and revised the manuscript.

## Code availability

All code to perform the analyses and generate the figures in this study are available from the corresponding author upon reasonable request.

