## Supplement for "Superspreading of SARS-CoV-2 infections: A Systematic Review and Meta-analysis"

**Running title.** Superspreading of SARS-CoV-2 infections

**Keywords**. COVID-19; SARS-CoV-2; superspreading; dispersion parameter; systematic review; meta-analysis.

**Table S1. Description of studies included in the systematic review and meta-analysis**

| **Study** | **Method** | **Dispersion parameter (*k*), (95% CI)** | **Period** | **Region** |
| --- | --- | --- | --- | --- |
| Sun K et al.[[1]](https://paperpile.com/c/24WLDq/HWkUq) | Negative binomial distribution | 0.30 (0.23, 0.39) | 2020-1-16 to 2020-4-3 | Mainland China |
| Adam DC et al.[[2]](https://paperpile.com/c/24WLDq/aBjrT) | Negative binomial distribution | 0.43 (0.29, 0.67) | 2020-1-23 to 2020-4-28 | Hong Kong,  China |
| Bi Q et al.[[3]](https://paperpile.com/c/24WLDq/6Y34W) | Negative binomial distribution | 0.58 (0.35,1.18) | 2020-1-14 to 2020-2-12 | Mainland China |
| He D et al.[[4]](https://paperpile.com/c/24WLDq/6iiy7) | Negative binomial distribution | 0.70 (0.59, 0.98) | 2020-1-15 to 2020-2-29 | Mainland China |
| Hasan A et al.[[5]](https://paperpile.com/c/24WLDq/9kOE7) | Negative binomial distribution | 0.06 (0.05, 0.07) | 2020-3-2 to 2020-3-31 | Indonesia |
| Hasan A et al.[[5]](https://paperpile.com/c/24WLDq/9kOE7) | Negative binomial distribution | 0.20 (0.09, 0.31) | 2020-3-19 to 2020-4-7 | Indonesia |
| Kwok KO et al.[[6]](https://paperpile.com/c/24WLDq/z8SrY) | Negative binomial distribution | 2.30 (0.02, 4.58) | By 2020-3-3 | Hong Kong,  China |
| Kwok KO et al.[[6]](https://paperpile.com/c/24WLDq/z8SrY) | Negative binomial distribution | 0.51 (0.21, 1.59) | By 2020-3-3 | Japan |
| Kwok KO et al.[[6]](https://paperpile.com/c/24WLDq/z8SrY) | Negative binomial distribution | 1.78 (0.09, 3.47) | By 2020-3-3 | **Singapore** |
| Lau MSY et al.[[7]](https://paperpile.com/c/24WLDq/YTVFK) | Negative binomial distribution | 0.63 (0.54, 0.85) | 2020-3-1 to 2020-4-3 | USA |
| Lau MSY et al.[[7]](https://paperpile.com/c/24WLDq/YTVFK) | Negative binomial distribution | 0.66 (0.60, 0.71) | 2020-3-1 to 2020-4-3 | USA |
| Lau MSY et al.[[7]](https://paperpile.com/c/24WLDq/YTVFK) | Negative binomial distribution | 0.62 (0.54, 0.75) | 2020-3-1 to 2020-4-3 | USA |
| Lau MSY et al.[[7]](https://paperpile.com/c/24WLDq/YTVFK) | Negative binomial distribution | 0.64 (0.53, 0.75) | 2020-3-1 to 2020-4-3 | USA |
| Lau MSY et al.[[7]](https://paperpile.com/c/24WLDq/YTVFK) | Negative binomial distribution | 0.39 (0.37, 0.44) | 2020-3-1 to 2020-4-3 | USA |
| Miller D et al.[[8]](https://paperpile.com/c/24WLDq/ehNyN) | Phylodynamic analysis | 2.97 (2.86, 3.08) | By 2020-4-22 | Israel |
| Tariq A et al. [[9]](https://paperpile.com/c/24WLDq/BW69O) | Negative binomial distribution | 0.11 (0.05, 0.25) | 2020-1-23 to 2020-3-17 | Singapore |
| Wang L et al.[[10]](https://paperpile.com/c/24WLDq/d91HA) | Phylodynamic analysis | 0.23 (0.13, 0.38) | 2019-12-24 to 2020-2-14 | Mainland China |
| Zhao S et al.[[11]](https://paperpile.com/c/24WLDq/qaJAi) | Negative binomial distribution (Zero-truncated framework) | 0.37 (0.29, 0.48) | 2020-1-15 to 2020-2-29 | Mainland China |
| Zhao S et al.[[11]](https://paperpile.com/c/24WLDq/qaJAi) | Negative binomial distribution (Zero-truncated framework) | 0.32 (0.15, 0.64) | 2020-1-23 to 2020-4-28 | Hong Kong, China |
| Zhao S et al.[[11]](https://paperpile.com/c/24WLDq/qaJAi) | Negative binomial distribution (Zero-truncated framework) | 0.18 (0.01, 1.79) | 2020-1-21 to 2020-2-26 | Mainland China |
| Zhang Y et al.[[12]](https://paperpile.com/c/24WLDq/EudDD) | Negative binomial distribution | 0.25 (0.13, 0.88) | 2020-1-21 to 2020-2-26 | Mainland China |
| Shi Q et al.[[13]](https://paperpile.com/c/24WLDq/bmJ5M) | Negative binomial distribution | 0.21 (0.13, 0.33) | 2020-1-21 to 2020-4-10 | Mainland China |
| James A et al.[[14]](https://paperpile.com/c/24WLDq/IfjBo) | Negative binomial distribution | 0.29 (0.10, 2.05) | 2020-3-25 to 2020-4-22 | New Zealand |
| Kremer C et al.[[15]](https://paperpile.com/c/24WLDq/tfrVU) | Negative binomial distribution | 0.43 (0.38, 0.49) | 2020-1-23 to 2020-4-18 | Hong Kong, China |
| Kremer C et al.[[15]](https://paperpile.com/c/24WLDq/tfrVU) | Negative binomial distribution | 0.50 (0.50, 0.51) | By 2020-8-1 | India |
| Kremer C et al.[[15]](https://paperpile.com/c/24WLDq/tfrVU) | Negative binomial distribution | 0.56 (0.29, 0.83) | By 2020-12-31 | Rwanda |
| Endo A.[[16]](https://paperpile.com/c/24WLDq/P3BuM) | Negative binomial distribution | 0.10 (0.05, 0.20) | By 2020-2-27 | Global |
| Riou J et al.[[17]](https://paperpile.com/c/24WLDq/T5DKl) | Negative binomial distribution | 0.54 (0.01, 8.18) | By 2020-1-18 | Global |

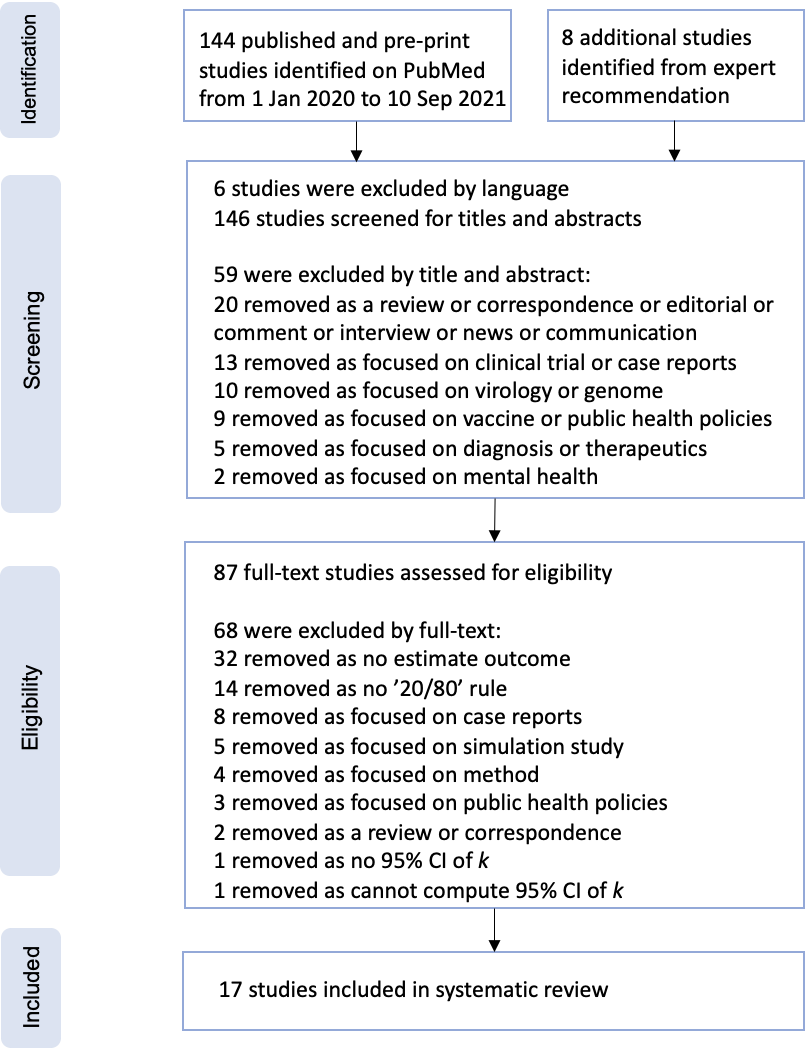

**Figure S1. PRISMA (Preferred Reporting Items for Systematic Reviews and Meta-Analyses) flow diagram for the studies used to obtain studies that reported measurements of the dispersion parameter.** We used PubMed for our primary search.

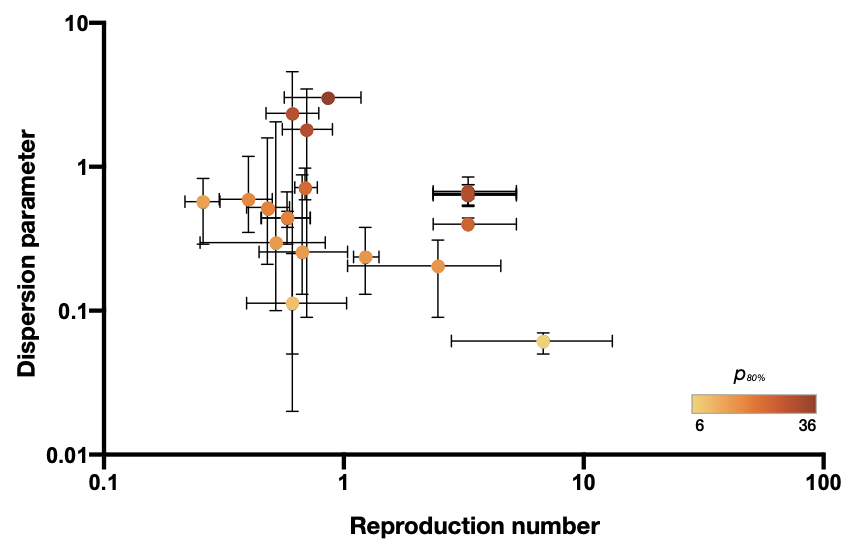

**Figure S2. Dispersion parameter estimates and reproduction numbers for coronavirus disease 2019 (COVID-19) reported in 19 unique studies presented by country.** The error bars show the mean values and 95% confidence interval of the dispersion parameter estimates and reported reproduction numbers in studies (**Supplement**). The color denotes the estimated proportion of cases accounting for 80% of all transmissions (*p_80%_*).

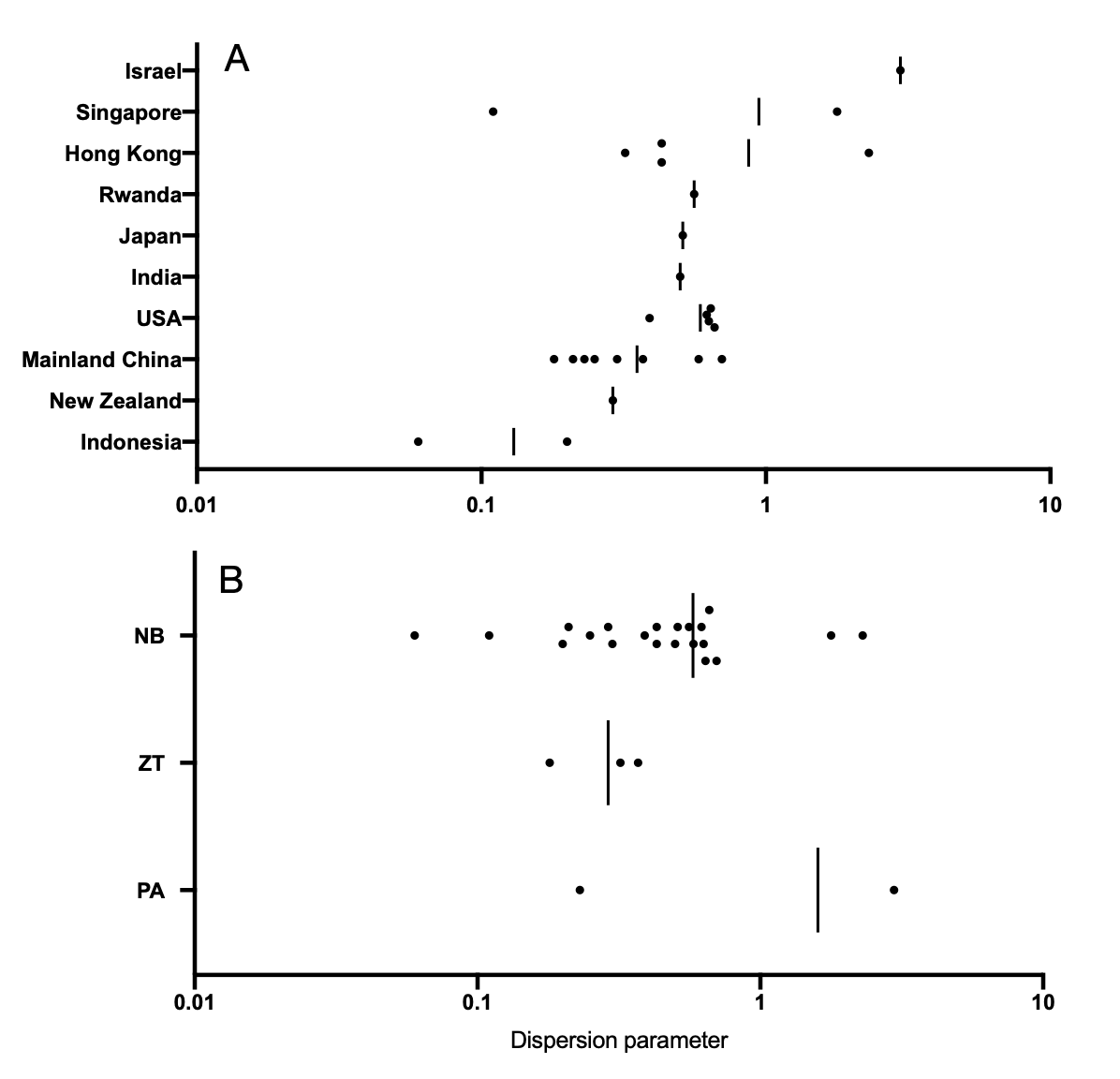

**Figure S3. Distribution of the estimated mean dispersion parameter with respect to (A) countries studied and (B) methods studied.** Black circles denote the mean estimates across studies. Vertical lines denote the mean values by averaging that for each country or method. NB: Negative binomial distribution; ZT: Negative binomial distribution (Zero-truncated framework); PA: Phylodynamic analysis.
